# Ceftazidime–Avibactam Resistance in *Escherichia coli* Primarily Mediated by *bla*_NDM-5_ with Emergent Ceftazidime–Avibactam/Aztreonam Resistance Linked to *bla*_CMY-42_ Variants

**DOI:** 10.64898/2025.12.05.25341718

**Authors:** William C Shropshire, Jovan Borjan, Ayesha Khan, Allyson Young, Jovelle K Chung, Chin-Ting Wu, Ambrocio Manzanares, Micah M Bhatti, Nancy N Vuong, Guy Handley, Amy Spallone, Roy F. Chemaly, Samuel Shelburne

## Abstract

Ceftazidime–avibactam (CZA) has revolutionized care for carbapenem-resistant Enterobacterales infections, yet increasing New Delhi metallo-β-lactamase (NDM) prevalence has driven use of CZA plus aztreonam (CZA/ATM). We performed a comprehensive analysis of CZA-resistant *Escherichia coli* (CZA-R-*Ec*) at a tertiary cancer center (2017–2024) by integrating clinical data, comparative genomics, and CZA/ATM susceptibility testing. CZA-R-*Ec* were isolated from forty-eight unique patients of whom 28 (58%) had confirmed infection. Oxford Nanopore Technologies long-read sequencing performed on 34 isolates from unique patients showed a diverse population enriched for ST167 (35%). Most sequenced isolates carried *bla*_NDM-5_ (26/34, 76%); among *bla*_NDM-5_ strains, 88% (23/26) harbored PBP3 YRI(K/N) insertions. Eleven isolates (32%) carried *bla*_CMY_ variants, predominantly *bla*_CMY-42_. B*la*_NDM-5_ and *bla*_CMY_ genes were largely plasmid-borne (IncF-type and IncI-γ/K1) in distinct genomic contexts. Among 32 confirmed CZA-R and ATM-R index isolates, 21 (66%) were CZA/ATM synergy positive to CZA/ATM (MIC≤4 µg/mL; SYN+). Compared to patients with SYN+ strains, patients with SYN-isolates (CZA/ATM MIC>4 µg/mL) were significantly more likely to have had a prior *E. coli* infection (73% vs. 0%, *P*-value=1.6×10^-5^). SYN- isolates were strongly associated with *bla*_CMY_ carriage (81% vs 9% in SYN+; adjusted *P*-value=4×10^-4^). Among 18 confirmed CZA-R-*Ec* bacteremias, 15 carried *bla*_NDM_; 14/15 (92%) received CZA/ATM and all responded clinically. In conclusion, CZA-R-*Ec* at our center are dominated by PBP3-insertion, *bla*_NDM-5_–positive lineages, for which CZA/ATM retains substantial *in vitro* activity and clinical efficacy. However, recent carbapenem exposure and prior *E. coli* infection identifies patients at risk for CZA/ATM non-synergy, frequently linked to *bla*_CMY-42_ variant positive isolates.

## INTRODUCTION

The introduction of ceftazidime/avibactam (CZA) along with other novel β-lactam/β-lactamase inhibitor combinations (BL/BLIs) has offered improved treatment options for carbapenem resistant Enterobacterales (CRE) infections [1–3]. Avibactam broadly inhibits Class A, C, and D β-lactamases, restoring ceftazidime activity against diverse Gram-negative organisms, including carbapenemase producers [1, 2]. However, avibactam does not inhibit metallo-β-lactamases (MBL) such as the New Delhi metallo-β-lactamase (NDM).

Therefore, in the context of MBL positive infections, clinicians have utilized CZA in combination with aztreonam (ATM), which relies on the concept that ATM is not hydrolyzed by MBLs while avibactam can inhibit many non-MBL ATM hydrolyzing enzymes (*e.g.*, *bla*_CTX-M_, *bla*_CMY_, and *bla*_OXA_) often co-carried in MBL producing strains. *In vitro* data support the synergistic efficacy of this combination [4], with fractional inhibitory concentration indices (FICI) <0.5 reported against various carbapenemase-producing strains [5, 6]. While large randomized controlled trials have evaluated the efficacy of CZA in treating Gram-negative infections broadly [7, 8], clinical data focused on MBL-producing *E. coli* and their association with ceftazidime-avibactam plus aztreonam (CZA/ATM) remain limited; in particular, most available evidence comes from case reports or small case series documenting microbiological eradication following CZA/ATM therapy [9–12]. Moreover, there are limited studies on the clinical effectiveness of CZA/ATM, particularly in immunocompromised hosts [11].

Notably, the *bla*_NDM-5_ MBL gene variant is being detected in multi-drug resistant (MDR) *E. coli* infections with increasing frequency worldwide [9, 13–19]. Whole genome sequencing (WGS) studies have identified several high-risk, MDR *E. coli* multilocus sequence types (MLSTs) associated with *bla*_NDM-5_ carriage, particularly ST167, ST101, ST410, ST361, and ST405, with a varied, worldwide geographic distribution [13–15, 17–20]. Many of these high-risk lineages possess fixed mutations in *ftsI*, encoding penicillin-binding protein 3 (PBP3), a principal β-lactam target, resulting in reduced affinity for agents such as ATM [17, 19, 21, 22]. These PBP3 mutations, in particular stable, four amino acid insertions of YRIN or YRIK following residue P333, have been associated with compromised *in vitro* CZA/ATM activity in clinical NDM producing pathogens, which further complicates treatment modalities [19, 20, 22, 23]. Simner *et al.* reported a case in which an insertion sequence (*i.e.*, IS*26*) based pseudocompound transposon (PCTN) was associated with increased copy number and expression of the *bla*_NDM-5_ gene with corresponding elevations in CZA/ATM and cefiderocol (FDC) minimum inhibitory concentrations (MICs) showing how gene dosage in the context of these *ftsI* insertion backgrounds can lead to pan-resistant infections [24]. Furthermore, we and others have recently detected high-risk *E. coli* lineages harboring plasmid mediated AmpC genes, in particular *bla*_CMY-2_ variants often in the presence of these *ftsI* mutations, which have high CZA MICs as well as reduced CZA/ATM activity [25–28].

While the aforementioned studies have described the genetic mechanisms underlying CZA resistance in *E. coli* and reported clinical management of CZA-R infections, few have integrated detailed genomic and microbiologic findings with clinical correlates. To address this gap, we conducted a comprehensive analysis of CZA-R *E. coli* (CZA-R-*Ec*) isolates at our institution, combining WGS, microbiologic characterization, and patient-level clinical data to elucidate the molecular mechanisms driving CZA and CZA/ATM resistance and subsequent impact on therapeutic response and clinical outcomes.

## RESULTS

### There has been a gradual increase in absolute frequency of annual CZA-R-*Ec* infections at our institution

There were 48 unique patients from whom CZA-R-*Ec* were isolated from 2017 to 2024. At our institution, no active screening for *E. coli* is performed so all strains were isolated as part of routine clinical care. **Table 1** provides a descriptive overview of these patients. The absolute frequency of CZA-R-*Ec* isolated from patients was 18 cases detected between 2017-2020 compared to 30 cases from 2021-2024. Many patients (n=23) were neutropenic (ANC < 500 cells/mm^3^) at the time of index CZA-R-*Ec* isolation, and 27% (13/48) were hematopoietic stem cell transplant (HSCT) recipients. A total of 58% (28/48) CZA-R-*Ec* cases were confirmed infections with the majority being bloodstream infections (68%, n=19). The most common treatment modality was CZA/ATM (74%;20/27) with one patient mortality occurring prior to initiation of treatment. The overall 30-day mortality following index CZA-R-*Ec* collection was 13% (6/48), with four deaths occurring in patients with confirmed CZA-R-*Ec* infection.

**Table 1:** Clinical characteristics of 48 patients from which ceftazidime-avibactam resistant Escherichia coli were isolated.

|  |  |
| --- | --- |
| Covariate | N, % unless specified |
| Age, median (min, max) | 63 (18, 88) |
| Gender, male | 28 (58) |
| International travel in previous 12 months | 25 (52) |
| Neutrophil count < 500 cells/mm<sup>3</sup> | 23 (48) |
| Hematopoietic Stem Cell Transplant Recipient | 13 (27) |
| Infection type (n= 28) |  |
| Blood | 19 (68) |
| Urine | 3 (11) |
| Sputum | 2 (7) |
| Other | 4 (14) |
| Treatment (n = 27) |  |
| CZA/ATM | 20 (74) |
| FDC | 3 (11) |
| Other | 4 (14) |
| 30-day mortality | 6 (13) |

### CZA-R-*Ec* primarily consist of high-risk lineages harboring *bla*_NDM-5_ and *ftsI* insertions

There were 34 unique patient isolates available for sequencing. A full list of genomic data for these plus recurrent isolates (n=7) sequenced can be found in Table S1. Clinical data comparison from patients with sequenced (n=34) vs. non-sequenced (n=14) isolates showed that sequenced isolates were more likely to have been infections (71% vs. 36%; *P*-value =0.05) and isolated from blood (53% vs 14%; *P*-value =0.03), but there was no significant difference amongst phenotypic carbapenemase testing results (80% vs. 86%; *P*-value: 1). **Fig. 1** provides an overview of the population structure as well as genomic features associated with CZA-R-*Ec* collected isolates. The most commonly detected sequence type (ST) was ST167 (35%; n=12), with other high-risk lineages such as ST361 (15%; n=5), ST405 (9%, n=4), ST410 (6%, n=3), and ST44 (6%, n=2) being observed more than once. Using a core genome, recombination masked alignment, we identified two clusters of closely related CZA-R-*Ec* isolates (*i.e.*, single nucleotide polymorphism (SNP) pairwise distances < 15 [29]) labelled Group 1 (ST167; n=5) and Group 2 (ST44; n=2) respectively (Fig. S1; Table S1), suggesting potential transmission. Group 1 consisted of *bla*_NDM-5_ positive ST167 isolates harboring p.P333insYRIN *ftsI* mutations whereas Group 2 consisted of two *bla*_NDM-1_ positive ST44 isolates.

**Fig 1.**
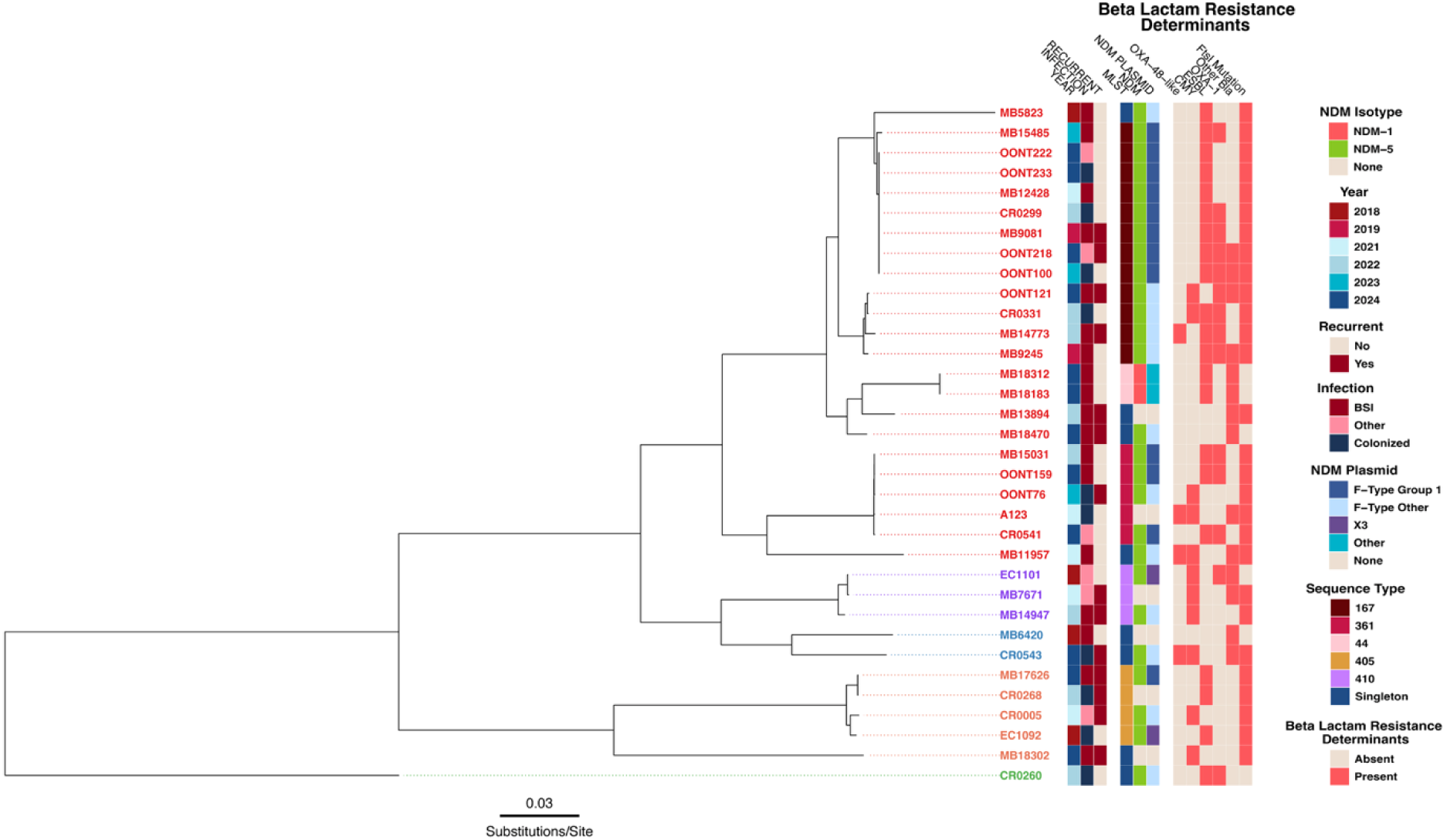
Core genome alignment maximum-likelihood inferred phylogeny of first occurrent CZA-R-*Ec* isolates (N=34) collected from 2018 to 2024. Terminal branch tips are colored based off phylogroup with phylogroup A (red), C (purple), B1 (blue), D (orange), and B2 (green) observed from top to bottom. Metadata from the heatmap is labelled accordingly in the legend present to the right of the phylogeny.

The majority (76%; 26/34) of the CZA-R-*Ec* isolates were *bla*_NDM-5_ positive with only two isolates (MB18183 and MB18312) harboring *bla*_NDM-1_ genes. Most of the *bla*_NDM-5_ positive isolates (88%; 23/26) had an *ftsI* mutation (17 with p.P333insYRIN and six with p.P333insYRIK). With the exception of one MBL positive isolate (MB18470), all MBL positive strains carried a combination of *bla*_CTX-M_ (*bla*_CTX-M-15_ [n=13], *bla*_CTX-M-14_ [n=2], *bla*_CTX-M-14_/ *bla*_CTX-M-15_ [n=1]), *bla*_OXA-1_ (n=14), *bla*_CMY_ (*bla*_CMY-42_ [n=5], *bla*_CMY-4_ [n=1], *bla*_CMY-59_ [n=1], *bla*_CMY-145_ [n=1]), and/or *bla*_OXA-48-like_ (*bla*_OXA-48_ [n=1], *bla*_OXA-1181_ [n=1], *bla*_OXA-1207_ [n=1]) genes, which further highlights the MDR genotype of these pathogens (Table S1). *Bla*_CMY-42_ variants were present in three of the six strains that were *bla*_NDM_ negative (**Fig. 1**).

### Integration of clinical and genomic data for CZA-R-*Ec* causing bloodstream infections

To integrate clinical and genomic data, we focused on the 18 patients with CZA-R*-Ec* bloodstream infections with sequencing data available, all of whom had hematologic malignancies and an absolute neutrophil count (ANC) < 500 cells/mm^3^. *Bla*_NDM_ was present in 15 of the 18 isolates. Among the three patients infected with MBL-negative strains, two died within 30 days. The third patient with an MBL-negative strain died 60 days after receiving sequential therapy with FDC, CZA/ATM plus tigecycline, and meropenem–vaborbactam plus eravacycline. Of the 15 patients with *bla*_NDM_ isolates, all but one were treated with CZA/ATM and had a favorable clinical response. The remaining patient received meropenem–vaborbactam, guided by clinical microbiology laboratory susceptibility results, and also responded well. Four of the 15 patients with *bla*_NDM_ -positive bloodstream infections experienced recurrent bacteremia, and three of these patients were successfully re-treated with β-lactam/β-lactamase inhibitor plus ATM combinations.

### CZA/ATM *in vitro* synergy was observed in majority of CZA-R-*Ec* isolates

Antimicrobial susceptibility (AST) results are shown in Table S2 for the 34 index and 7 recurrent isolates that tested initially CZA-R based off electronic health records. We confirmed CZA-R phenotypes in all but one of our 34 available isolates (MB6420), which tested CZA-S upon repeat testing (Table S2). This index bloodstream infection isolate has truncated *ompC* and *ompF* genes with concomitant increased *bla*_TEM-1_ gene copy numbers (∼150 copies); these gene copy numbers can be unstable and could account for the loss of CZA-R phenotype [29–32]. We further excluded an additional isolate that tested CZA-R but ATM-S (MB18470) from our CZA/ATM analysis with this phenotype likely due to the absence of β-lactam resistance mechanisms other than *bla*_NDM-5_ and *bla*_TEM-1_.

**Fig. 2** provides clinical and genomic details for the first available CZA-R and aztreonam resistant (ATM-R) isolates upon which we performed CZA/ATM MIC testing. Among 32 confirmed CZA-R and ATM-R-*Ec* index isolates tested with reference broth microdilution, 21 (66%) had CZA/ATM MICs <=4 μg/mL (for convenience purposes, we refer to these isolates as SYN+ and isolates with CZA/ATM MICS >4 μg/mL as SYN-). Of the 11 SYN- index isolates, eight (73%) were from patients who had at least one *E. coli* infection in the 100 days prior to the index CZA-R-*Ec* isolate (**Fig.2**; **Table 2**) whereas none of the patients with SYN+ isolates had a previous *E. coli* infection within this timeframe (*P*-value=1.6×10^-5^). A comparable proportion of patients had previous cephalosporin exposure within 100 days from index CZA-R-*Ec* isolate for those with SYN- (8/11;73%) vs SYN+ (17/21; 81%; adjusted *P*-value=0.3) strains. However, a higher proportion of patients with SYN- isolates had previous carbapenem exposure in the 100 days prior to CZA-R-*Ec* isolation (9/11; 82%) compared to SYN+ isolates (7/21; 33%) (**Table 2**; adjusted *P*-value=0.05). The median days of any cephalosporin exposure 100 days prior in the SYN+ group (6 days; IQR: 9 days) was similar in duration to the SYN- group (4 days; IQR: 7 days) (adjusted *P*-value=0.4). The median days of any carbapenem exposure in the 100 days prior was greater in the SYN- group (12 days; IQR: 13 days) compared to SYN+ group (0 days; IQR: 7 days) (adjusted *P*-value=0.02).

**Fig 2.**
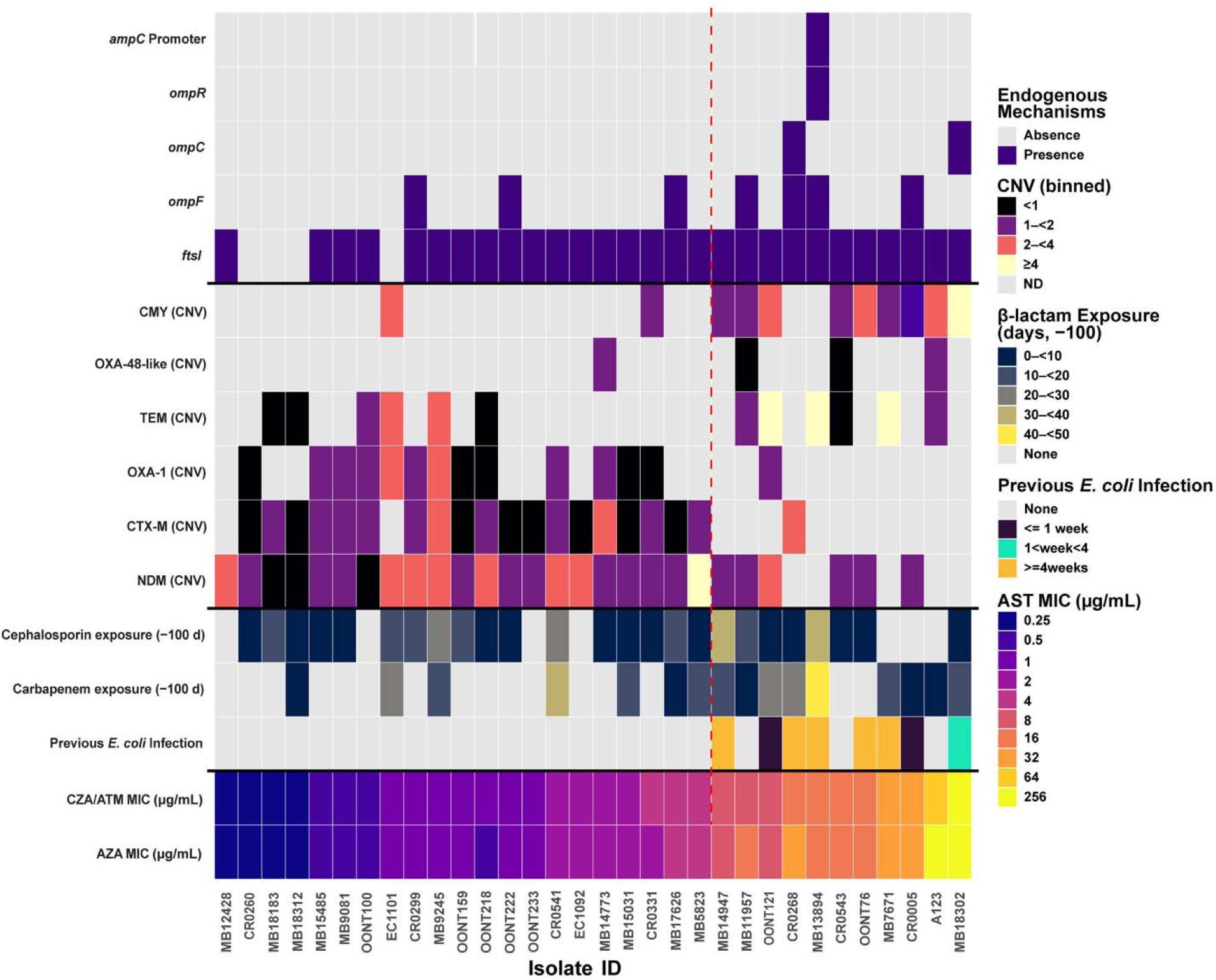
Clinical and genomic features associated with increasing CZA/ATM and AZA MICs. The x-axis is sorted first by increasing CZA/ATM MIC then by increasing ATM and CZA MIC respectively. Y-axis from top to bottom indicates: Rows 1-5: presence/absence of chromosomal mutations predicted to impact B-lactam resistance; Rows 6-11: presence/absence and copy number variation (CNV) of exogenous B-lactamase content; Rows 12-14: Clinical metadata. Vertical dotted red line indicates cutoff between CZA/ATM SYN+ (left) and CZA/ATM SYN-(right). Metadata from the heatmap is labelled accordingly in the legend present to the right of the figure.

**Table 2:**
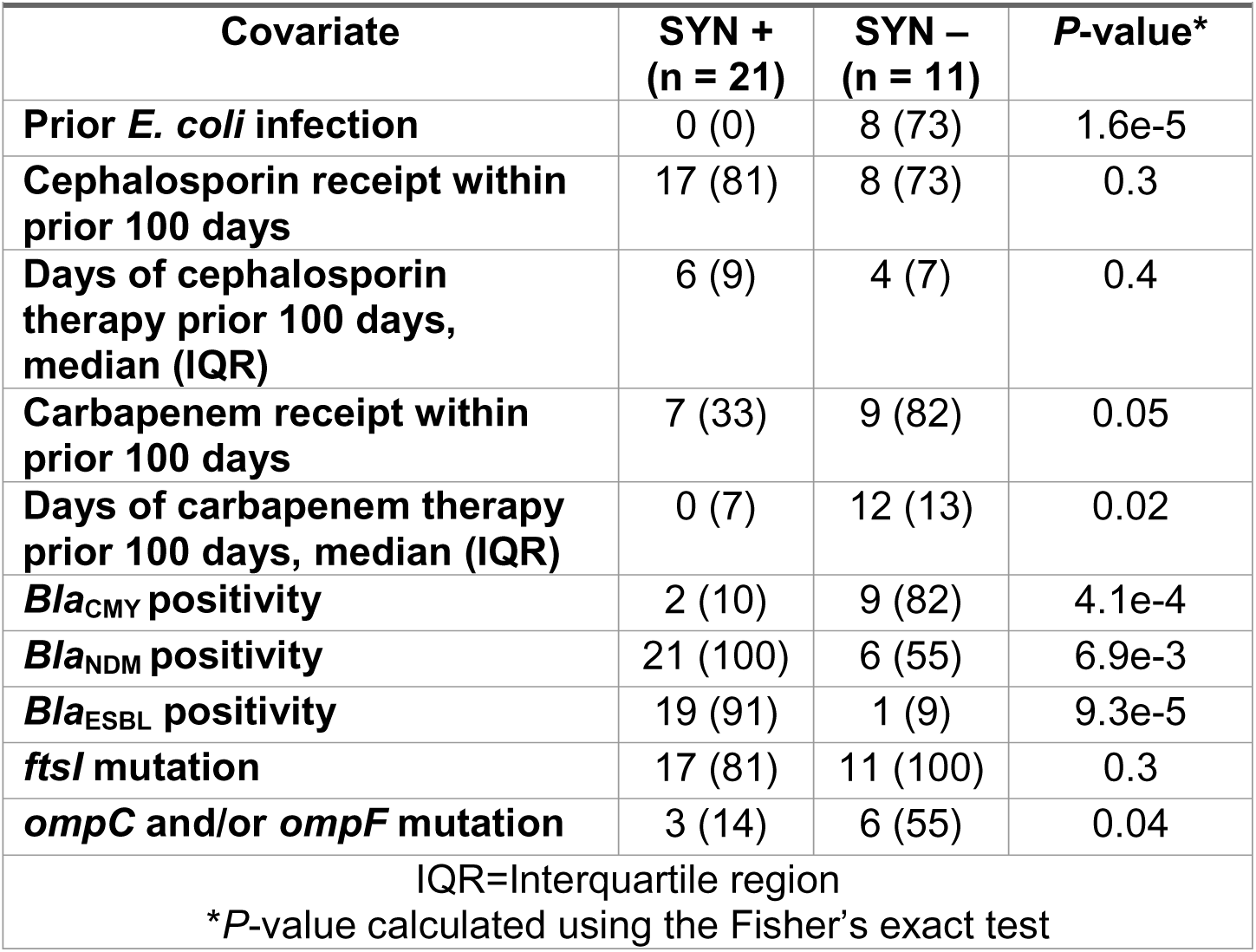
Comparison of ceftazidime/avibactam + aztreonam SYN + vs. SYN- strains.

|  |  |  |  |
| --- | --- | --- | --- |
| Covariate | SYN +<br>(n = 21) | SYN –<br>(n = 11) | P-value* |
| Prior *E. coli* infection | 0 (0) | 8 (73) | 1.6e-5 |
| Cephalosporin receipt within prior 100 days | 17 (81) | 8 (73) | 0.3 |
| Days of cephalosporin therapy prior 100 days, median (IQR) | 6 (9) | 4 (7) | 0.4 |
| Carbapenem receipt within prior 100 days | 7 (33) | 9 (82) | 0.05 |
| Days of carbapenem therapy prior 100 days, median (IQR) | 0 (7) | 12 (13) | 0.02 |
| *Bla*<sub>CMY</sub> positivity | 2 (10) | 9 (82) | 4.1e-4 |
| *Bla*<sub>NDM</sub> positivity | 21 (100) | 6 (55) | 6.9e-3 |
| *Bla*<sub>ESBL</sub> positivity | 19 (91) | 1 (9) | 9.3e-5 |
| *ftsI* mutation | 17 (81) | 11 (100) | 0.3 |
| *ompC* and/or *ompF* mutation | 3 (14) | 6 (55) | 0.04 |
| IQR=Interquartile region |  |  |  |
| *P-value calculated using the Fisher's exact test |  |  |  |

A notable observation was the higher proportion of SYN- isolates that carried *bla*_CMY_ (9/11; 82%) in contrast to SYN+ isolates (2/23; 10%) (adjusted *P*-value: 4.1×10^-4^) (**Fig.2**; **Table 2**). Stratifying by *bla*_CMY-42-like_ variants (*i.e.*, CMY alleles with or without p.V221 mutations relative to CMY-2), all SYN- isolates had a p.V221 mutation with two isolates also harboring p.N346 mutations (MB7671 and A123). *Bla_CMY_* variants detected by SYN classification with additional genomic context data (*e.g.*, other *bla* genes, copy numbers) can be found in Table S3. MB7671 has been previously described [25, 26] as having four mutations relative to CMY-2 (Ambler positions A114E, Q120K, V211S, N346Y). Kawai *et al.* established p.N346Y as the main driver of CMY-185 mediated CZA resistance by both blocking avibactam binding and enhancing ceftazidime hydrolysis, while V211S further boosts resistance by increasing the turnover rate of oxyimino-cephalosporin hydrolysis [26]. Strain A123 carries a novel *bla*_CMY_ gene, designated *bla*_CMY-225_, which has three mutations relative to CMY-2 (Ambler positions N70T, V211S, and N346D) and a concomitantly high CZA/ATM MIC (64 μg/mL; **Fig. 2**) in a PBP3 p.P333insYRIN background. The two SNY+ strains which encoded *bla*_CMY_ included one isolate with a wild type *ftsI* background (EC1101) and another (CR0331) which contained *bla*_CMY-4_ (*i.e.*, non *bla*_CMY-42-like_) variant.

**Figure 2** and **Table 2** also highlight additional genomic features such as PBP3 predicted YRI(K/N) duplication mutations being present in all SYN- isolates and most SYN+ (17/21, 81%) isolates (adjusted *P*-value=0.3). There was a statistically significant higher CZA/ATM MIC in PBP3 YRI(K/N) positive isolates (3 μg/mL; IQR: 15 μg/mL) compared to PBP3 YRI(K/N) negative isolates (0.25 μg/mL; IQR: 0.19 μg/mL) (adjusted P-value=0.02). Carriage of *bla*_NDM_, *bla*_CTX-M_, and *bla*_OXA-1_ was more common in SYN+ isolates (100%, 91%, and 62% respectively) compared to SYN- isolates (55%, 9%, and 9% respectively) (**Table 2**; adjusted *P*-values < 0.02). The two SYN-/*bla*_CMY_- isolates lacked *bla*_NDM_ as well and instead carried disparate genomic backgrounds. One isolate (CR0268) was *ompC/ompF*-deficient and carried a predicted PBP3 p.P333insYRIK mutation with *bla*_CTX-M-15_. The other (MB13894) harbored *ompF* and *ompR* disrupting mutations along with *ampC* promoter mutations in addition to *bla*_TEM-1_. When testing differences in copy number variants (CNVs) of *bla*_NDM-5_, there was no discernable difference in SYN+ (mean 1.9x; SD: 1.2) and SYN- (mean 1.6x; SD: 0.36; *P*-value=0.7) strains.

Finally, we compared the distribution of CZA/ATM with aztreonam-avibactam (AZA) MICs (**Fig 2.,** Table S2). With the exception of one isolate (A123; CZA/ATM MIC=64 μg/mL; AZA MIC=256 μg/mL), all others were within one doubling dilution of CZA/ATM and AZA MICs, and no statistically significant difference was detected after doing a log2 MIC transformation (Wilcoxon on log2 MICs: *P*-value=0.2). Furthermore, when comparing results from CZA/ATM reference broth microdilution and the qualitative ETEST-based strip cross method [30], we found perfect concordance when calling CZA/ATM MICs <=4 μg/mL vs > 4 μg/mL, reaffirming the practicality and reliability of the latter method to detect SYN- CZA-R*-Ec* strains in the clinical setting (Table S2).

### *Bla*_NDM-5_ and *bla*_CMY_ variants predominantly carried on highly conserved F-Type and IncI-γ/K1 plasmids

With the exception of a single patient (CR0005/CR0063 isolates) in whom *bla*_CMY_ occurred in a chromosomal rather than plasmid context, all *bla*_CMY_ and *bla*_NDM_ genes were plasmid-borne. In addition, nearly all confirmed CZA-R-*Ec* isolates (30/32) harbored *bla*_NDM_ and/or *bla*_CMY_; thus, we performed a comprehensive comparative plasmid analysis to (1) determine how these AMR genes clustered with previously characterized complete plasmids available on NCBI, (2) assess whether β-lactamase gene copy number was driven by plasmid copy number or by mobile genetic elements (MGEs), and (3) identify the associated mobilization mechanisms.

**Fig. 3A** shows that most *bla*_NDM-5/-1_ plasmids in our combined cohort and public dataset cluster within IncF subcommunities; IncF subtypes predominantly carry *bla*_NDM-5_, whereas *bla*_NDM-1_ is chiefly associated with IncC and IncN subcommunities (Fig. S2A). Similar to our strains, we found that 20% of *bla*_NDM_ positive plasmids originated from ST167 strains (147/724) with many of the IncF *bla*_NDM-5_ positive isolates having been collected in the USA and Europe in the past four years (Fig. S2B-C; Table S4).

**Fig 3.**
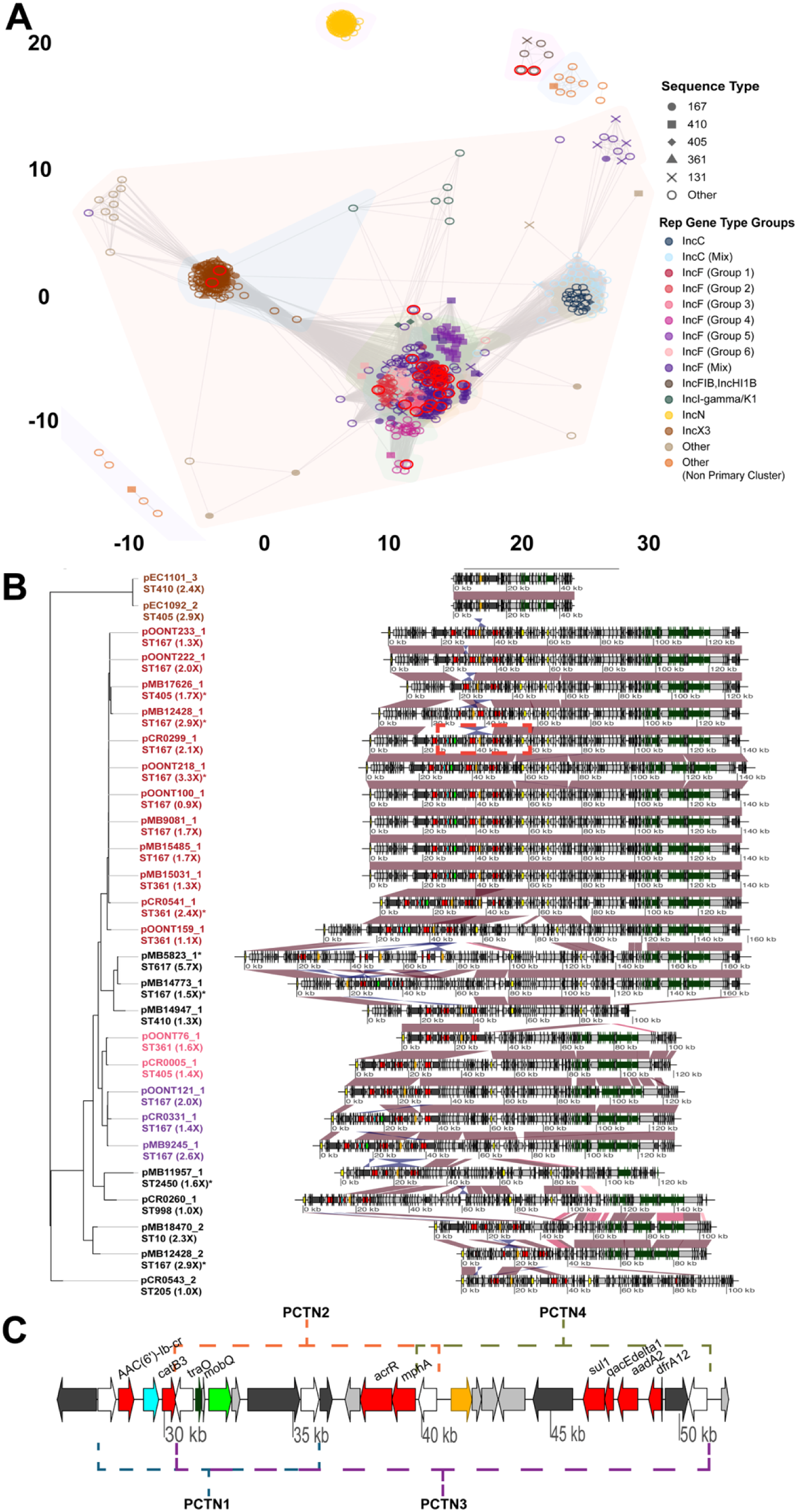
Plasmids harboring *bla*_NDM_ comparative genomics analysis. **(A)** Network analysis of full-length plasmids harboring *bla*_NDM-1_ and *bla*_NDM-5_ identified in our study (red circles around shapes) and NCBI data abstracted using Pathogen Detection. Shapes indicate most commonly identified sequence types and color represent Inc type groupings based off pling clustering results. Subcommunity linkage is indicated by gray edges connecting nodes (*i.e.*, plasmid sequences). Peripheral groups (*e.g.*, IncN plasmids) share no linkage with primary community identified in center of figure. **(B)** Blastn comparison of full-length multireplicon plasmids. Branch tips are colored by groupings established in subcommunity analysis with sequence type labeled. Black font indicates a ‘singleton’ plasmid that doesn’t group with another plasmid in our cohort. Number in parenthetical represents estimated *bla*_NDM-5_ copy number from AMR-STRUCT with asterisk indicating increase in *bla*_NDM-5_ copy number was associated with PCTN amplification. Red and blue shading between sequences indicates 95% shared blastn ID in direct and reverse orientation respectively. *Bla*_NDM-5_ (orange), *bla*_CTX-M-15_ (green), *bla*_OXA-1_ (blue), other AMR genes (red), MGE content (dark grey), rep genes (yellow), IS6 family transposase gene (white), Tn3 genes (brown), and conjugal transfer genes (dark green) are labelled accordingly. Plasmids reoriented to IncF replication initiation gene (yellow) for DNA comparisons. Dotted region indicates zoomed region in following subsection. **(C)** Zoomed in region of F-Type plasmid (Group 1; 25844 – 51913 bp – pCR0299_1 reference) with well conserved PCTNs detected in Group 1 plasmids.

Focusing on our *bla*_NDM-5_ plasmids, nearly half (12/27) belonged to a predicted conjugative IncFIA,IncFIB,IncFIC,IncFII multireplicon plasmid cluster (IncF Group 1) which shares megablast weighted averages of >99% identity and 95% coverage (**Fig. 3B**). The cargo gene region for this plasmid cluster includes a well conserved, mosaic ∼25kb pseudocompound transposon (PCTN) that carries *bla*_NDM-5_ in addition to *bla*_CTX-M-15_ and *bla*_OXA-1_ with four predicted IS*26* PCTN units (**Fig. 3C**) based on previous studies of IS*26* mobilization mechanisms [31]. Using AMR-STRUCT (See methods), four of the 12 IncF Group 1 plasmids had predicted PCTN increased copies based off increase in coverage depths within PCTN region relative to the total plasmid coverage depth or 2 or more resolved copies in the complete assembly (**Fig. 3C**). Three amplified units are PCTN4 detected on pCR0541_1, pOONT218_1, and pMB17626_1 with two copies of PCTN4 fully resolved on pCR0541_1 (**Fig. 2B**); The PCTN harboring *bla*_NDM-5_ on pMB12428_1 is a novel 15.8kb PCTN that has an additional copy on pMB12428_2 present on a unique IncFIA,IncFIC 92.2kb plasmid.

**Fig. 4A** provides an overview of the network of *bla*_CMY_ positive plasmids abstracted from NCBI in addition to those from our own cohort. The majority of *bla*_CMY_ positive plasmids (n=310) belonged to IncI-γ/K1 based subcommunities (71%; 219/310) with two other groups (IncC/IncK2 based subcommunities) also present (**Fig. 4A**). When stratifying the IncI-γ/K1 subcommunities by *bla*_CMY_ variant, Group 1 carried predominantly *bla*_CMY-42_ or *bla*_CMY-42_ like variants (70%; 132/188), in particular, variants with the V211S amino acid substitution, whereas Group 2 carried *bla*_CMY-2_ (96%; 22/23) (**Fig. 4A**, Fig S3).

**Fig 4.**
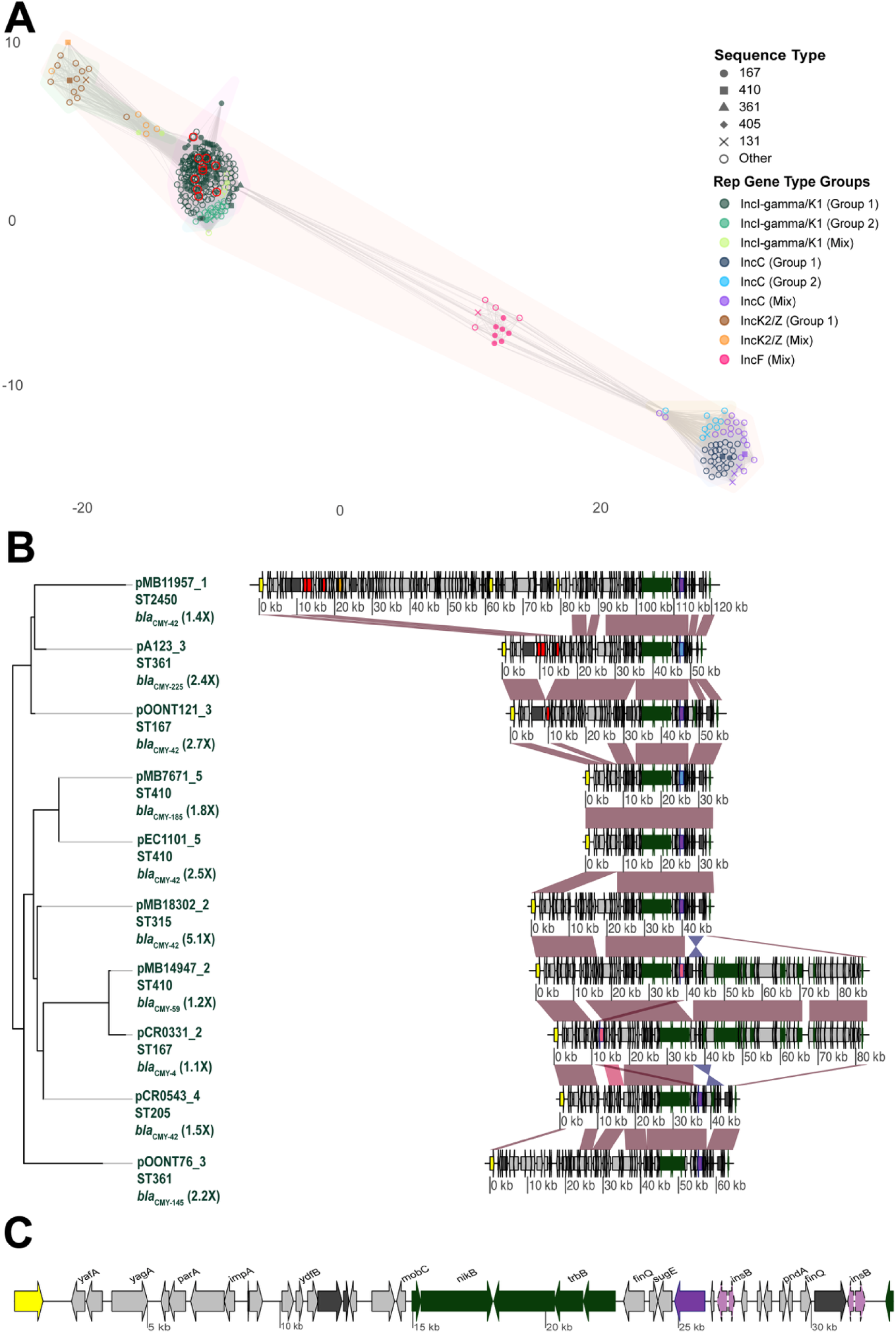
Plasmids harboring *bla*_CMY_ comparative genomics analysis. **(A)** Network analysis of full-length plasmids harboring *bla*_CMY-2_ and *bla*_CMY-2_ variants identified in our study (red circles around shapes) and NCBI data abstracted using Pathogen Detection. Shapes indicate most commonly identified sequence types and color represent Inc type groupings based off pling clustering results. Subcommunity linkage is indicated by gray edges connecting nodes (*i.e.*, plasmid sequences). **(B)** Blastn comparison of full-length IncI-gamma/K1 plasmids. All plasmids belonged to group 1 as represented in **(A)**. Number in parenthetical represents estimated *bla*_CMY_ copy number from AMR-STRUCT. Red and blue shading between sequences indicates 95% shared blastn ID in direct and reverse orientation respectively. *Bla*_NDM-5_ (orange), other AMR genes (red), MGE content (dark grey), rep genes (yellow), IS6 family transposase gene (white), Tn3 genes (brown), and conjugal transfer genes (dark green) are labelled accordingly. Non V211S blaCMY-2 variants are pink, V211S *bla*_CMY-2_ variants are labelled in purple, and V211S *bla*_CMY-2_ variants with p.N346 mutations are labelled in blue. **(C)** Representative IncI-gamma/K1 plasmid (pEC1101 reference) with IS1R genes upstream labelled in light pink and dotted outline stroke.

All 10 of the *bla*_CMY_ plasmids in our cohort belonged to the Group 1 IncI-γ/K1 subcommunity which shared a mean 98.6% megablast identity and 71% coverage (**Fig. 4B**). Seven of the ten plasmids had *bla*_CMY_ estimated copy numbers >1.5× with all seven estimated to be due to plasmid copy number changes rather than transposon mediated mobilization (**Fig. 4B**). **Fig. 4C** illustrates a representative IncI-γ/K1 plasmid (pEC1101_5) with the highly conserved gene order of transposase IS1R (*insB*) upstream of *bla*_CMY_ shaded in purple/blue dependent on *bla*_CMY_ variant type (**Fig. 4B**). Interestingly, pMB7671_5 and pEC1101_5 shared >99.9% blast identity, 100% coverage and estimated plasmid copy number >1.5×, with the notable difference that pMB7671_5 harbored the inhibitor resistant *bla*_CMY-185_ variant whereas pEC1101_5 harbored a *bla*_CMY-42_ variant (**Fig. 4B**). Only one plasmid (pMB11957_1) co-carried *bla*_NDM-5_ and *bla*_CMY-42_ in our cohort (**Fig. 4B**), which was a recombined IncI-γ/K1 and IncFIB/IncFII plasmid (Table S5). This low plasmid co-carriage was consistent with our global CMY plasmid analysis where only 2% (5/310) of *bla*_CMY_ positive plasmids carried *bla*_NDM-5_ (Fig. S3; Table S5).

### Recurrent CZA-R-*Ec* isolates share similar CZA/ATM phenotypes as index isolates

We had seven patients with index CZA-R-*Ec* and recurrent CZA-R-*Ec* isolates available for sequencing (Table S6). The median time from index to recurrent isolate collection for these pairs was 113 days (range: 49 – 569 days). All paired isolates were collected from infections (5 bloodstream, 2 other) and median number of recurrences was 2 (range: 1 – 5). Two patients (Patient 2 and Patient 5) died within 25 and 6 days respectively of their final recurrent CZA-R-*Ec* bacteremia. When performing comparative genomics on index to recurrent isolate, the median pairwise SNP distance was 3 (range: 2-6) (Table S6).

Among the 7 patients with paired index and recurrent CZA-R-*Ec* isolates, two pairs were classified as SYN- with the remaining 5 patients having SYN+ paired isolates (Table S6). The only sequential isolates with a substantial CZA/ATM and/or AZA MIC change (*i.e.*, change of greater/less than 1 log2 fold dilution MIC for recurrent isolate) were from patient 2 (pairwise SNP distance: 6), which were also the only *bla*_NDM-5_ negative recurrent isolates. Genomic changes for these isolates included a predicted p.Leu369Phe mutation in PBP3 (i.e., c.1107A>T in *ftsI*), as well as mutations in the efflux RND transporter permease AcrB (p.Phe136Leu) and envelope stress sensor histidine kinase CpxA (p.Gln120Lys) (Table S7). We also observed a *bla*_NDM-5_, *bla*_OXA-1_, and *bla*_CTX-M-15_ CNV increase in Patient 5 from 1.5x to 4.5x (Fig. S4) which was found to map to a 20kb unique PCTN as presented in **Fig. 3B** and Fig. S4; however, there was only a 2-fold increase in CZA/ATM and AZA MICs (Tables S2).

## DISCUSSION

Over the past 15 years, most advances in β-lactam therapy of drug-resistant Gram-negative bacteria have centered on new BL/BLI combinatorial treatments. Novel agents such as CZA have improved outcomes, particularly for infections caused by class A carbapenemase producers. However, the global expansion of MBL producers now threatens the utility of these agents. In this study, we integrated clinical and genomic analyses of CZA-R-*Ec* and found that CZA-R was predominantly driven by *bla*_NDM-5_, whereas reduced activity of the CZA/ATM combination was largely associated with carriage of *bla*_CMY-42_ and related variants.

Because many NDM-producing isolates come from non-sterile sites (*e.g.*, urine, sputum), clinical response can be hard to attribute to antimicrobial therapy. Our cohort’s high rate of bacteremia in profoundly immunocompromised patients offered a robust study frame to evaluate antimicrobial efficacy and link outcomes with genomics and detailed susceptibility profiles. We found that patients with CZA-R-*Ec* bacteremia carrying *bla*_NDM-5_ generally responded well to CZA/ATM, and that these outcomes aligned with *in vitro* results by reference broth microdilution and gradient cross-strip testing. These findings are consistent with recent reports demonstrating strong CZA/ATM activity against NDM-producing bloodstream infections, although one study was dominated by *Klebsiella pneumoniae* [12] and another included only four CZA-R-*Ec* treated with CZA/ATM monotherapy [11].

A second key finding was the clear clinical and genomic distinction between synergy-positive (SYN+) and synergy-negative (SYN−) CZA/ATM phenotypes. SYN+ isolates typically co-carried *bla*_NDM_ with *bla*_CTX-M-15_/*bla*_OXA-1_ in PBP3 YRI(K/N) insertion backgrounds and occurred in patients with less carbapenem exposure and no recent *E. coli* infection. In contrast, SYN− isolates tended to be isolated from patients with recent receipt of carbapenems and prior *E. coli* infection and were enriched for *bla*_CMY-42_ variants in PBP3 YRI(K/N) insertion backgrounds with frequent predicted OmpC/OmpF loss-of-function. These findings are consistent with reports in *E. coli* where CMY-42, PBP3 insertions, and porin defects act together (*i.e.*, through increased β-lactam hydrolysis, reduced target engagement, and lowered periplasmic drug levels) to reduce AZA susceptibility [20, 32–36]. Clinically, our data suggest that in CMY-42/PBP3-insertion backgrounds, prior carbapenem exposure alone may select for CZA (and AZA) non-susceptibility, even without prior CZA or ATM use. Importantly, our data also suggest that CMY-42 severely impacts the utility of CZA/ATM and AZA in NDM-producing isolates with PBP3 insertions that would otherwise be susceptible to these agents, limiting therapeutic options for clinicians. As AZA becomes more widely available in the U.S., the strong concordance we observed between CZA/ATM and AZA MICs supports using caution when empirically choosing AZA for CZA-R strains with this clinical and/or genomic risk profile. Additionally, our strong MIC concordance for CZA/ATM and AZA, including via CZA/ATM qualitative ETEST-based strip cross method, is a practical finding for clinical microbiology labs, lending confidence to use CZA/ATM testing as a surrogate for AZA susceptibility until non-research use only (RUO) commercial testing is widely available and validated.

Our long-read WGS resolved the genetic context of β-lactamase gene loci, allowing us to clearly differentiate *bla*_NDM_- and *bla*_CMY_-harboring plasmids. Our findings were consistent with other studies that detected *bla*_NDM-5_ in multireplicon IncF and to a lesser degree IncX3 genomic contexts in disparate geographical locales [24, 37, 38]. In our study, 82% of CZA-R-*Ec* carried *bla*_NDM-5_, typically on multireplicon hybrid F-type plasmids, in particular, one particular subcommunity we designated F-Type Group 1 that was commonly detected in ST167 isolates collected in the US and Europe. Our plasmids frequently carried *bla*_NDM-5_ within IS*26*-mediated PCTNs, consistent with mechanisms that both amplify *bla*_NDM-5_ copy number, which is associated with increased CZA and CZA/ATM MICs, and promote mobilization [24]. Notably, *bla*_NDM-5_ plasmids were found almost exclusively in strains with PBP3 insertions, echoing global observations that the majority of *E. coli* with PBP3 insertions harbor *bla*_NDM_, usually *bla*_NDM-5_ [39]. In our cohort, the *bla*_CMY_ gene was exclusively found in IncI-γ/K1 plasmid backgrounds with increased plasmid copy number inferred as likely mechanism for increase in *bla*_CMY_ gene dosage as inferred by long-read pileup coverage depths. IncI complex plasmids have been shown to be a common vector for *bla*_CMY-2_ transmission [40]; however, recent studies have shown that *bla*_CMY-2_ variants that develop inhibitor resistance associated with p.N346 mutations have also been associated with this plasmid type [25, 28, 41].

Although PBP3 insertions in an NDM-5 background increase MICs to CZA/ATM and AZA [20], we observed that CZA/ATM retained both *in vitro* activity and favorable clinical response in strains with PBP3 insertions provided *bla*_CMY-42_ like variants were absent. Because CMY variants are often missing from routine AMR gene panels, these findings support confirmatory phenotypic testing of CZA/ATM or AZA activity for NDM-positive cases before finalizing therapy. Our single-center, observational design and modest sample size are limitations, but integrating detailed clinical data with long-read genomics allowed clear separation of synergy-positive and - negative lineages. As NDM-5 continues to expand globally [14], larger, multi-center studies are needed to define optimal empiric strategies and to determine how antimicrobial exposure, plasmid context, and mobile elements shape treatment response and the emergence of CZA/ATM or AZA non-susceptibility. Our findings also highlight *bla*_CMY-42_ and related variants, whether or not *bla*_NDM_ is present, as priority targets for continued surveillance, mechanistic study, and directed therapy development.

## MATERIALS AND METHODS

### Study Design

CZA-R-*Ec* infections that occurred at our institution from 2017-01-01 to 2024-11-30 were abstracted from Epic using a custom workbench reporting tool. Clinical data and available susceptibility testing were abstracted from Epic via chart review. The MDACC IRB panel designated this study exempt from review (OHRP IRB Registration Number: IRB00000121). Recurrent CZA-R-*Ec* is defined as one or more episodes of CZA-R-*Ec* infection or colonization occurring at least 14 days following the initial CZA-R-*Ec* infection or colonization isolate while maintaining CZA-R phenotype with infection defined using standard CDC criteria [42]. Prior *E. coli* infection was considered any *E. coli* CZA-S infection that was detected prior to the initial CZA-R-*Ec* infection or colonization within 100 days from that index collection date. Prior carbapenem exposure was defined as one or more days of either ertapenem, meropenem, and/or imipenem therapy within 100 days prior from the index CZA-R-*Ec* infection or colonization date. Prior cephalosporin exposure was defined as one or more days of third/fourth generation cephalosporin therapy (*e.g.*, ceftriaxone, cefepime, and/or ceftazidime) within 100 days prior from the index CZA-R-*Ec* infection or colonization date. Exposure was counted as days of therapy (DOT) for each antibiotic which is a National Healthcare Safety Network (NHSN) metric for tracking antimicrobial use [43].

### Antimicrobial Susceptibility Testing

For clinical care purposes, antimicrobial susceptibility testing was primarily done with Vitek 2 (bioMerieux, Marcy-l’Étoile, France) with phenotypic carbapenemase testing performed using the Neo-Rapid CARB Kit (Rosco Diagnostica, Albertslund, Denmark) or RAPIDEC CARBA NP Assay (bioMérieux, Marcy-l’Étoile, France). CZA/ATM and AZA susceptibility testing for all available isolates was performed using standard broth microdilution assays as described [30, 44]. Gradient cross-strip testing was performed for CZA/ATM MTS (Liofilchem, Italy) and ATM E-test (bioMérieux, Marcy-l’Étoile, France) and in combination using previously established methodologies [44].

### Oxford Nanopore Technologies (ONT) long-read sequencing

There was a total of 12 CZA-R-*Ec* isolates that had existing ONT data from previous projects [25, 29, 45]. An additional 22 available first occurrent CZA-R-*Ec* isolates and seven recurrent CZA-R-*Ec* isolates that were saved in our −80°C stocks were subject to ONT sequencing. Genomic DNA was extracted using the Sigma-Aldrich GenElute Bacterial Genomic DNA Kit per manufacturer’s instructions. The gDNA was checked for concentration and quality using the Thermo Fisher Scientific Qubit 1X dsDNA Broad Range Assay kit and Agilent Genomic DNA SceenTape Analysis kit. ONT Library prep was performed using the ONT Rapid Barcoding Kit 96 V14 (SQK-RBK114.96) and R10.4.1 flowcells on a MinION Mk1D sequencing device. Basecalling was performed using a custom python script (run_dorado.py: https://github.com/wshropshire/misc_scripts) on the MDACC Seadragon high performance computing (HPC) cluster with the GPU enabled Dorado (v0.8.2) basecaller using a super accurate model (<u>dna_r10.4.1_e8.2_400bps_sup@v5.0.0</u>). Assemblies were created using a Flye assembler workflow (https://github.com/wshropshire/flyest) and manually checked and corrected for errors [46]. The DNA Variant Identification using ONT (dviont-v0.2.1), which incorporates clair3 (v1.1.2) [47] variant calling using the r1041_e82_400bps_sup_v430_bacteria_finetuned model, was utilized to identify potential errors (https://github.com/wshropshire/dviont). CheckM2-v1.1.0 was used for assembly quality control with all assemblies having 100% completeness and <1% contamination [48].

### Phylogenomics

Thirty-four first occurrent CZA-R-*Ec* isolates were selected to determine population structure of our cohort. Mash distances and FastANI were run with complete assemblies to determine which isolate had the lowest mean genetic distance from the total cohort to use as a reference for core genome alignment (reference=CR0299) [49, 50]. Parsnp (v2.1.1) was used to perform the core genome alignment with the muscle aligner and recombination filter parameters [51]. The core SNP alignment file was then used to infer a maximum-likelihood phylogeny using iqtree2 (v2.3.6) with ModelFinder (TVM+F+ASC+R7), ultrafast bootstrap approximation (n=1000), and SH-like aLRT branch support testing (n=1000) options [52, 53]. Phylogenetic tree with metadata heatmap visualization was implemented using the R ‘ggtree’ package (v.3.10.1) [54]. The core SNP alignment file generated from Parnsp (v2.1.1) was used to create a pairwise SNP distance matrix using snp-dists (v0.8.2) (https://github.com/tseemann/snp-dists). Subsequently, we constructed a minimum spanning tree (MST) of index ESC-R-*Ec* isolates, clustered using a SNP distance threshold of 15 based off previous literature [55, 56], and visualized with GraphSNP (v1.1) [57].

### blaNDM and blaCMY harboring plasmid cluster analysis

We extracted *bla*_CMY_ and *bla*_NDM-_5/*bla*NDM_-1_ positive sequences from NCBI (Accessed 2025-10) by using the Pathogen Detection Isolate Browser function to pull *E. coli* assemblies with (1) complete Amrfinder hits for the aforementioned genes, (2) 4-6 Mb in total genome length and (3) less than 15 contigs. We used NCBI Datasets command-line interface (CLI) to retrieve assemblies and accompanying metadata respectively utilizing RefSeq accession numbers pulled from Pathogen Detection Isolate Browser (https://www.ncbi.nlm.nih.gov/pathogens/isolates). Quality control was performed for all sequences using checkM2 where all assemblies had less than 5% contamination and greater than 99% completeness [48]. We analyzed all extracted plasmid sequences (contigs less than 300kb) using the mob_typer CLI function from the mob-suite toolkit (v3.1.9) and applied quality control by excluding sequences that lacked evidence of primary clustering with plasmids in our database [58]. We finally clustered plasmids with Pling-v2.0.0, which first computes containment distances of plasmids and “integerises” sequences, then calculates “Double Cut and Join-Indel” (DCJ–Indel) distances and builds a plasmid network on which communities/subcommunities are called capturing both sequence variation in addition to structural rearrangements [59]. Pairwise DCJ–Indel distances from Pling (all_plasmids_distances.tsv) were converted into weighted edges to construct an undirected plasmid similarity network in *R* (igraph-v2.1.4; https://r.igraph.org/). Node metadata (replicon type, sequence type, resistance genes, and collection features) were overlaid, and network visualization used a Fruchterman–Reingold layout with convex hulls outlining Pling-defined communities (ggraph-v2.2.2/ggforce-v0.4.2). *bla*_NDM-5_ and *bla*_CMY_ plasmids from our cohort were compared all-v-all using blastn (length limit = 1000 bp). Pairwise plasmid distances were then estimated using mash-v2.3 [49] (k-mer size 21; sketch size 1000) and this dissimilarity matrix was used to construct a bionj tree [60]. Megablast was performed on IncF Group 1 and IncI-γ/K1 Group1 plasmids respectively to calculate weighted mean % identity and coverage. Plasmids were then visualized using the R genoPlotR (v0.8.11) package [61].

### Additional computational genomic analysis

AMR-STRUCT (v0.1.0) was used as a wrapper of AMRFinderplus (v.4.0.19) with the 2025-07-16.1 database to (1) identify AMR genes; (2) perform minimap2 alignments on chromosome and plasmids (i.e., contigs >=1 Mb and < 1Mb respectively); and (3) perform sliding-window median coverage depth calculations with chromosome/plasmid and identify which AMR genes were likely amplified (*i.e.*, >1.5× normalized coverage depth ratios) and in what context (*i.e.*, plasmid, MGE) (https://github.com/wshropshire/AMR-STRUCT). MGE predicted amplifications were confirmed by qualitatively determining if increased mapping at boundaries of predicted MGE using bam files and visualized on IGV (v.2.19.1). Mutations in outer membrane porin as well as penicillin binding protein encoding genes were detected using the Mutation Identification in Sequences and Frameshift/Indel Tracking (misfit-v0.0.1) with completed assemblies as input (https://github.com/wshropshire/misfit). A ONT long-read variant calling tool, the DNA Variant Identification using ONT (dviont-v0.2.1), which serves as a clair3 wrapper tool [47], was used to determine pairwise SNP distances and annotate mutations in serial isolates (https://github.com/wshropshire/dviont).

### Statistics

All statistics were computed on R-v4.5.1. Fisher’s exact test was used to test statistical significance of categorical variables whereas Wilcoxon rank-sum tests were used to test statistically significant differences of continuous variables. *P*-values were adjusted using the Benjamini-Hochberg procedure to control the false discovery rate in cases where multiple comparisons (*e.g.*, antibiotic usage, AMR gene/feature) were made.

### Data Availability

Genomic data and assemblies from prior publications are available under NCBI BioProjects PRJNA836696, PRJNA924946, and PRJNA1150149. Data generated for this study have been deposited under PRJNA1308153 with data details and BioSample Accession Numbers provided in Table S1. All other data produced in the present study are available upon reasonable request to the authors.

## ACKNOWLEDGMENTS

SAS received support for this study through NIAID R21AI151536 and P01AI152999.

## REFERENCES

1. Soriano, A., et al., Ceftazidime-Avibactam for the Treatment of Serious Gram-Negative Infections with Limited Treatment Options: A Systematic Literature Review. Infectious Diseases and Therapy, 2021. 10(4): p. 1989–2034.

2. Sharma, R., T.E. Park, and S. Moy, Ceftazidime-Avibactam: A Novel Cephalosporin/beta-Lactamase Inhibitor Combination for the Treatment of Resistant Gram-negative Organisms. Clin Ther, 2016. 38(3): p. 431–44.

3. Cruz-López, F., et al., Efficacy and In Vitro Activity of Novel Antibiotics for Infections With Carbapenem-Resistant Gram-Negative Pathogens. Frontiers in Cellular and Infection Microbiology, 2022. 12.

4. Wise, M.G., et al., In vitro activity of aztreonam–avibactam against Enterobacterales isolates collected in Latin America, Africa/Middle East, Asia, and Eurasia for the ATLAS Global Surveillance Program in 2019–2021. European Journal of Clinical Microbiology &amp; Infectious Diseases, 2023. 42(9): p. 1135–1143.

5. Boattini, M., et al., Activity of cefiderocol and synergy of novel β-lactam-β lactamase inhibitor-based combinations against metallo-β-lactamase-producing gram-negative bacilli: insights from a two-year study (2019–2020). Journal of Chemotherapy, 2023. 35(3): p. 198–204.

6. Lu, G., et al., In vitro and in vivo Antimicrobial Activities of Ceftazidime/Avibactam Alone or in Combination with Aztreonam Against Carbapenem-Resistant Enterobacterales. Infection and Drug Resistance, 2022. 15: p. 7107–7116.

7. Carmeli, Y., et al., Ceftazidime-avibactam or best available therapy in patients with ceftazidime-resistant Enterobacteriaceae and Pseudomonas aeruginosa complicated urinary tract infections or complicated intra-abdominal infections (REPRISE): a randomised, pathogen-directed. The Lancet Infectious Diseases, 2016. 16(6): p. 661–673.

8. Wagenlehner, F.M., et al., Ceftazidime-avibactam Versus Doripenem for the Treatment of Complicated Urinary Tract Infections, Including Acute Pyelonephritis: RECAPTURE, a Phase 3 Randomized Trial Program. Clinical Infectious Diseases, 2016. 63(6): p. 754–762.

9. Khan, A., et al., Simultaneous infection with Enterobacteriaceae and Pseudomonas aeruginosa harboring multiple carbapenemases in a returning traveler colonized with Candida auris. Antimicrobial agents and chemotherapy, 2020. 64(2): p. e01466–19.

10. Marshall, S., et al., Can Ceftazidime-Avibactam and Aztreonam Overcome βLactam Resistance Conferred by Metallo-β-Lactamases in Enterobacteriaceae? Antimicrobial Agents and Chemotherapy, 2017. 61(4): p. AAC.02243-16.

11. Wang, J., et al., A comparative analysis of clinical outcomes in hematological patients afflicted with bacteremia attributable to carbapenem-resistant Klebsiella pneumoniae versus Escherichia coli. Frontiers in Cellular and Infection Microbiology, 2025. 15.

12. Falcone, M., et al., Efficacy of Ceftazidime-avibactam Plus Aztreonam in Patients With Bloodstream Infections Caused by Metallo-β-lactamase–Producing Enterobacterales. Clinical Infectious Diseases, 2021. 72(11): p. 1871–1878.

13. Hans, J.B., et al., Molecular surveillance reveals the emergence and dissemination of NDM-5-producing Escherichia coli high-risk clones in Germany, 2013 to 2019. Eurosurveillance, 2023. 28(10).

14. Linkevicius, M., et al., Rapid cross-border emergence of NDM-5-producing Escherichia coli in the European Union/European Economic Area, 2012 to June 2022. Eurosurveillance, 2023. 28(19).

15. Sun, P., et al., Characterization of blaNDM-5-positive Escherichia coli prevalent in a university hospital in eastern China. Infection and drug resistance, 2019: p. 3029–3038.

16. Simner, P.J., et al., An NDM-Producing Escherichia coli Clinical Isolate Exhibiting Resistance to Cefiderocol and the Combination of Ceftazidime-Avibactam and Aztreonam: Another Step Toward Pan-beta-Lactam Resistance. Open Forum Infect Dis, 2023. 10(7): p. ofad276.

17. Tellapragada, C., et al., Resistance to aztreonam-avibactam among clinical isolates of Escherichia coli is primarily mediated by altered penicillin-binding protein 3 and impermeability. Int J Antimicrob Agents, 2024. 64(3): p. 107256.

18. Sadek, M., et al., International circulation of aztreonam/avibactam-resistant NDM-5-producing Escherichia coli isolates: successful epidemic clones. J Glob Antimicrob Resist, 2021. 27: p. 326–328.

19. Sadek, M., et al., Genetic Features Leading to Reduced Susceptibility to Aztreonam-Avibactam among Metallo-β-Lactamase-Producing Escherichia coli Isolates. Antimicrobial Agents and Chemotherapy, 2020. 64(12).

20. Livermore, D.M., et al., Activity of aztreonam/avibactam against metallo-beta lactamase-producing Enterobacterales from the UK: Impact of penicillin-binding protein-3 inserts and CMY-42 beta-lactamase in Escherichia coli. Int J Antimicrob Agents, 2023. 61(5): p. 106776.

21. Periasamy, H., et al., High prevalence of Escherichia coli clinical isolates in India harbouring four amino acid inserts in PBP3 adversely impacting activity of aztreonam/avibactam. Journal of Antimicrobial Chemotherapy, 2020. 75(6): p. 1650–1651.

22. Alm, R.A., M.R. Johnstone, and S.D. Lahiri, Characterization of Escherichia coli NDM isolates with decreased susceptibility to aztreonam/avibactam: role of a novel insertion in PBP3. Journal of Antimicrobial Chemotherapy, 2015. 70(5): p. 1420–1428.

23. Zhang, Y., et al., Unusual Escherichia coli PBP 3 Insertion Sequence Identified from a Collection of Carbapenem-Resistant Enterobacteriaceae Tested In Vitro with a Combination of Ceftazidime-, Ceftaroline-, or Aztreonam-Avibactam. Antimicrobial Agents and Chemotherapy, 2017. 61(8): p. e00389–17.

24. Simner, P.J., et al., Progressive Development of Cefiderocol Resistance in Escherichia coli During Therapy is Associated With an Increase in blaNDM-5 Copy Number and Gene Expression. Clinical Infectious Diseases, 2022. 75(1): p. 47–54.

25. Shropshire, W.C., et al., High-level ceftazidime/avibactam resistance in Escherichia coli conferred by the novel plasmid-mediated β-lactamase CMY-185 variant. Journal of Antimicrobial Chemotherapy, 2023. 78(10): p. 2442–2450.

26. Kawai, A., et al., Structural insights into the molecular mechanism of high-level ceftazidime-avibactam resistance conferred by CMY-185. mBio, 2024. 15(2): p. e0287423.

27. Xu, T., et al., Novel plasmid-mediated CMY variant (CMY-192) conferring ceftazidime-avibactam resistance in multidrug-resistant Escherichia coli. Antimicrobial Agents and Chemotherapy, 2024. 68(12).

28. Zhou, J., et al., A Novel CMY Variant Confers Transferable High-Level Resistance to Ceftazidime-Avibactam in Multidrug-Resistant Escherichia coli. Microbiology Spectrum, 2023. 11(2): p. e03349–22.

29. Wu, C.-T., et al., Rapid whole genome characterization of antimicrobial-resistant pathogens using long-read sequencing to identify potential healthcare transmission. Infection Control & Hospital Epidemiology, 2024. 46(2): p. 1–7.

30. Khan, A., et al., Evaluation of two gradient diffusion tests to determine susceptibility to aztreonam and ceftazidime–avibactam in combination. Antimicrobial Agents and Chemotherapy, 2025. 69(3).

31. Harmer, C.J. and R.M. Hall, IS26 and the IS26 family: versatile resistance gene movers and genome reorganizers. Microbiology and Molecular Biology Reviews, 2024. 88(2).

32. Deshpande, L.M., et al., Activity of aztreonam-avibactam and ceftazidime-avibactam against beta-lactamase-producing enterobacterales Isolates from United States hospitals. J Glob Antimicrob Resist, 2025. 44: p. 103–110.

33. Long, H., et al., Global distribution of blaCMY-42, a gene mediating reduced susceptibility to aztreonam-avibactam and ceftazidime-avibactam, in Escherichia coli. Int J Antimicrob Agents, 2024. 63(5): p. 107141.

34. Mushtaq, S., et al., Activity of aztreonam/avibactam and ceftazidime/avibactam against Enterobacterales with carbapenemase-independent carbapenem resistance. Int J Antimicrob Agents, 2024. 63(3): p. 107081.

35. Helsens, N., et al., Reduced susceptibility to aztreonam-avibactam conferred by acquired AmpC-type β-lactamases in PBP3-modified Escherichia coli. European Journal of Clinical Microbiology &amp; Infectious Diseases, 2024.

36. Ma, K., et al., Struggle To Survive: the Choir of Target Alteration, Hydrolyzing Enzyme, and Plasmid Expression as a Novel Aztreonam-Avibactam Resistance Mechanism. mSystems, 2020. 5(6).

37. Chakraborty, T., et al., Cross-Border Emergence of Escherichia coli Producing the Carbapenemase NDM-5 in Switzerland and Germany. Journal of Clinical Microbiology, 2021. 59(3).

38. Zou, H., et al., Emergence of NDM-5-Producing Escherichia coli in a Teaching Hospital in Chongqing, China: IncF-Type Plasmids May Contribute to the Prevalence of blaNDM–5. Frontiers in Microbiology, 2020. 11.

39. Long, H., et al., Global emergence of Escherichia coli with PBP3 insertions. Journal of Antimicrobial Chemotherapy, 2025. 80(1): p. 178–181.

40. Sidjabat, H.E., et al., Expansive spread of IncI1 plasmids carrying blaCMY-2 amongst Escherichia coli. Int J Antimicrob Agents, 2014. 44(3): p. 203–8.

41. Xu, M., et al., Emergence of transferable ceftazidime–avibactam resistance in KPC-producing Klebsiella pneumoniae due to a novel CMY AmpC β-lactamase in China. Clinical Microbiology and Infection, 2022. 28(1): p. 136.e1–136.e6

42. Prevention, U.S.C.f.D.C.a., National Healthcare Safety Network (NHSN) Patient Safety Component Manual. 2025, Division of Healthcare Quality Promotion Atlanta, GA.

43. Morris, A.M., Antimicrobial Stewardship Programs: Appropriate Measures and Metrics to Study their Impact. Current Treatment Options in Infectious Diseases, 2014. 6(2): p. 101–112.

44. Khan, A., et al., Evaluation of Susceptibility Testing Methods for Aztreonam and Ceftazidime-Avibactam Combination Therapy on Extensively Drug-Resistant Gram-Negative Organisms. Antimicrobial Agents and Chemotherapy, 2021. 65(11).

45. Shropshire, W., et al., Systematic Analysis of Mobile Genetic Elements Mediating β-Lactamase Gene Amplification in Noncarbapenemase-Producing Carbapenem-Resistant Enterobacterales Bloodstream Infections. mSystems, 2022. 7(5): p. e00476–22.

46. Kolmogorov, M., et al., Assembly of long, error-prone reads using repeat graphs. Nat Biotechnol, 2019. 37(5): p. 540–546.

47. Zheng, Z., et al., Symphonizing pileup and full-alignment for deep learning-based long-read variant calling. Nat Comput Sci, 2022. 2(12): p. 797–803.

48. Chklovski, A., et al., CheckM2: a rapid, scalable and accurate tool for assessing microbial genome quality using machine learning. Nature Methods, 2023. 20(8): p. 1203–1212.

49. Ondov, B.D., et al., Mash: fast genome and metagenome distance estimation using MinHash. Genome Biol, 2016. 17(1): p. 132.

50. Jain, C., et al., High throughput ANI analysis of 90K prokaryotic genomes reveals clear species boundaries. Nature Communications, 2018. 9(1).

51. Kille, B., et al., Parsnp 2.0: scalable core-genome alignment for massive microbial datasets. Bioinformatics, 2024. 40(5).

52. Minh, B.Q., et al., IQ-TREE 2: New Models and Efficient Methods for Phylogenetic Inference in the Genomic Era. Mol Biol Evol, 2020. 37(5): p. 1530–1534.

53. Kalyaanamoorthy, S., et al., ModelFinder: fast model selection for accurate phylogenetic estimates. Nature Methods, 2017. 14(6): p. 587–589.

54. Yu, G., et al., ggtree: package for visualization and annotation of phylogenetic trees with their covariates and other associated data. Methods in Ecology and Evolution, 2017. 8(1): p. 28–36.

55. Sundermann, A.J., et al., Whole-genome sequencing surveillance and machine learning for healthcare outbreak detection and investigation: A systematic review and summary. Antimicrob Steward Healthc Epidemiol, 2022. 2(1): p. e91.

56. Rebelo, A.R., et al., One day in Denmark: whole-genome sequence-based analysis of Escherichia coli isolates from clinical settings. Journal of Antimicrobial Chemotherapy, 2025. 80(4): p. 1011–1021.

57. Permana, B., S.A. Beatson, and B.M. Forde, GraphSNP: an interactive distance viewer for investigating outbreaks and transmission networks using a graph approach. BMC Bioinformatics, 2023. 24(1).

58. Robertson, J. and J.H.E. Nash, MOB-suite: software tools for clustering, reconstruction and typing of plasmids from draft assemblies. Microbial Genomics, 2018. 4(8).

59. Frolova, D., et al., Applying rearrangement distances to enable plasmid epidemiology with pling. Microb Genom, 2024. 10(10).

60. Gascuel, O., BIONJ: an improved version of the NJ algorithm based on a simple model of sequence data. Molecular Biology and Evolution, 1997. 14(7): p. 685–695.

61. Guy, L., J. Roat Kultima, and S.G.E. Andersson, genoPlotR: comparative gene and genome visualization in R. Bioinformatics, 2010. 26(18): p. 2334–2335.

