## Supplemental Figures for "Ceftazidime–Avibactam Resistance in *Escherichia coli* Primarily Mediated by *bla*_NDM-5_ with Emergent Ceftazidime–Avibactam/Aztreonam Resistance Linked to *bla*_CMY-42_ Variants"

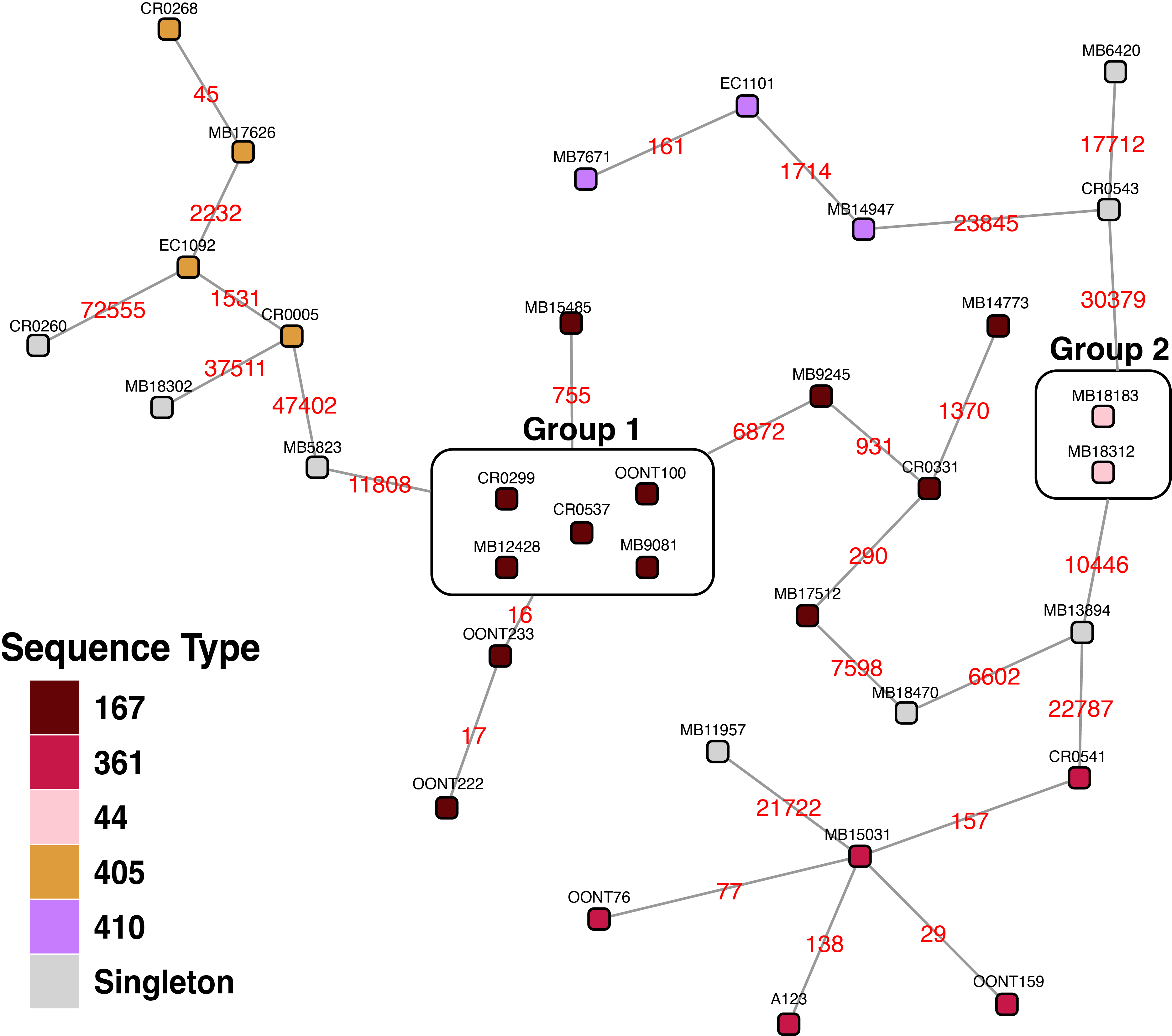

**Fig S1** Minimum spanning tree with nodes (isolates) connected by edges (pairwise SNP distances) with red text indicating pairwise SNP distance. Nodes are colored by sequence type with Group 1 ST167 and Group 2 ST44 isolates (<15 pairwise SNPs) circled and labelled respectively.

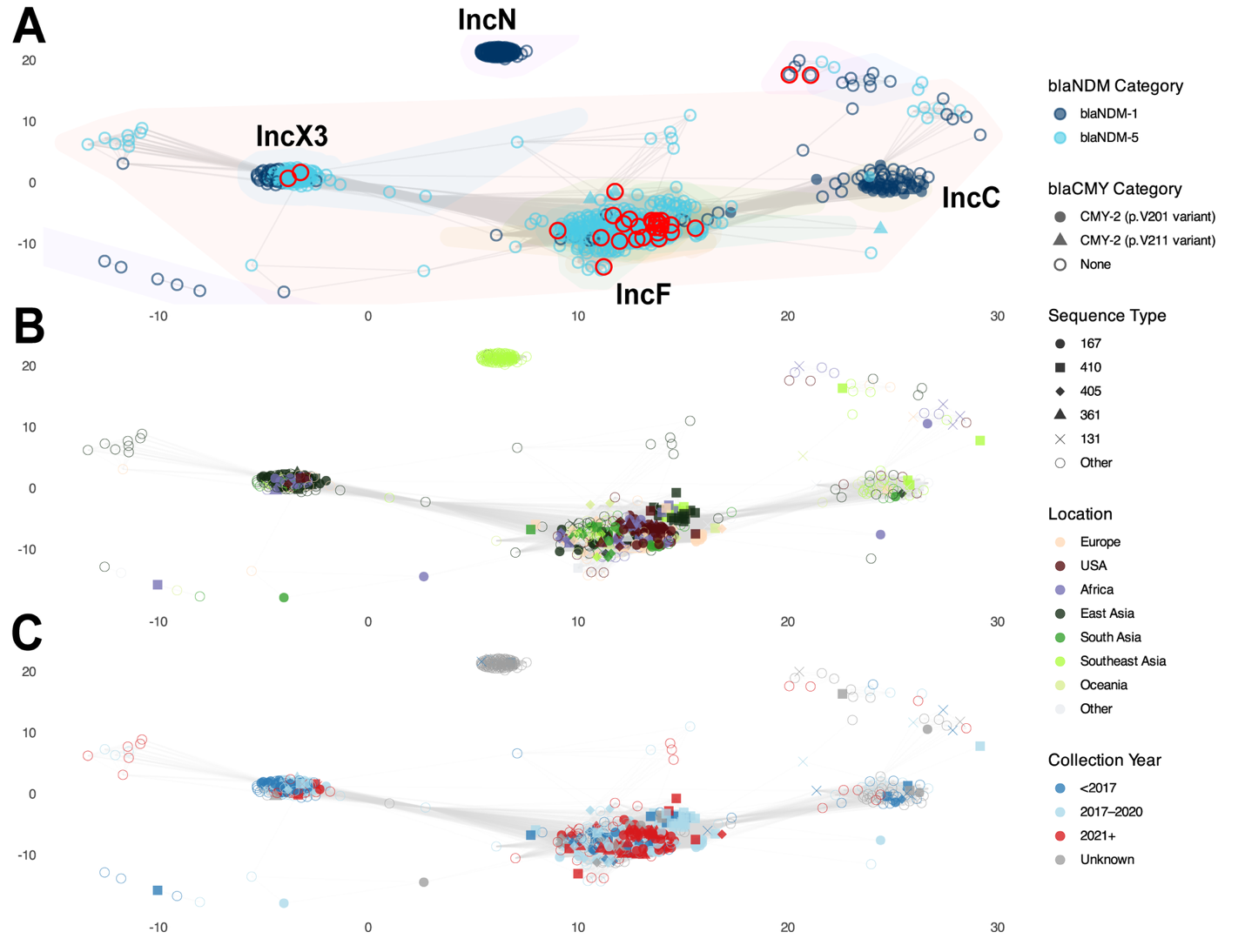

**Fig S2** NDM plasmid network analysis stratified by **(A)** *bla*_NDM_ variant carriage and *bla*_CMY_ variant carriage; **(B)** sequence type and location collected; **(C)** sequence type and collection year. Plasmid groups are labelled as well as red circles identifying this study plasmid cohort in panel **A**.

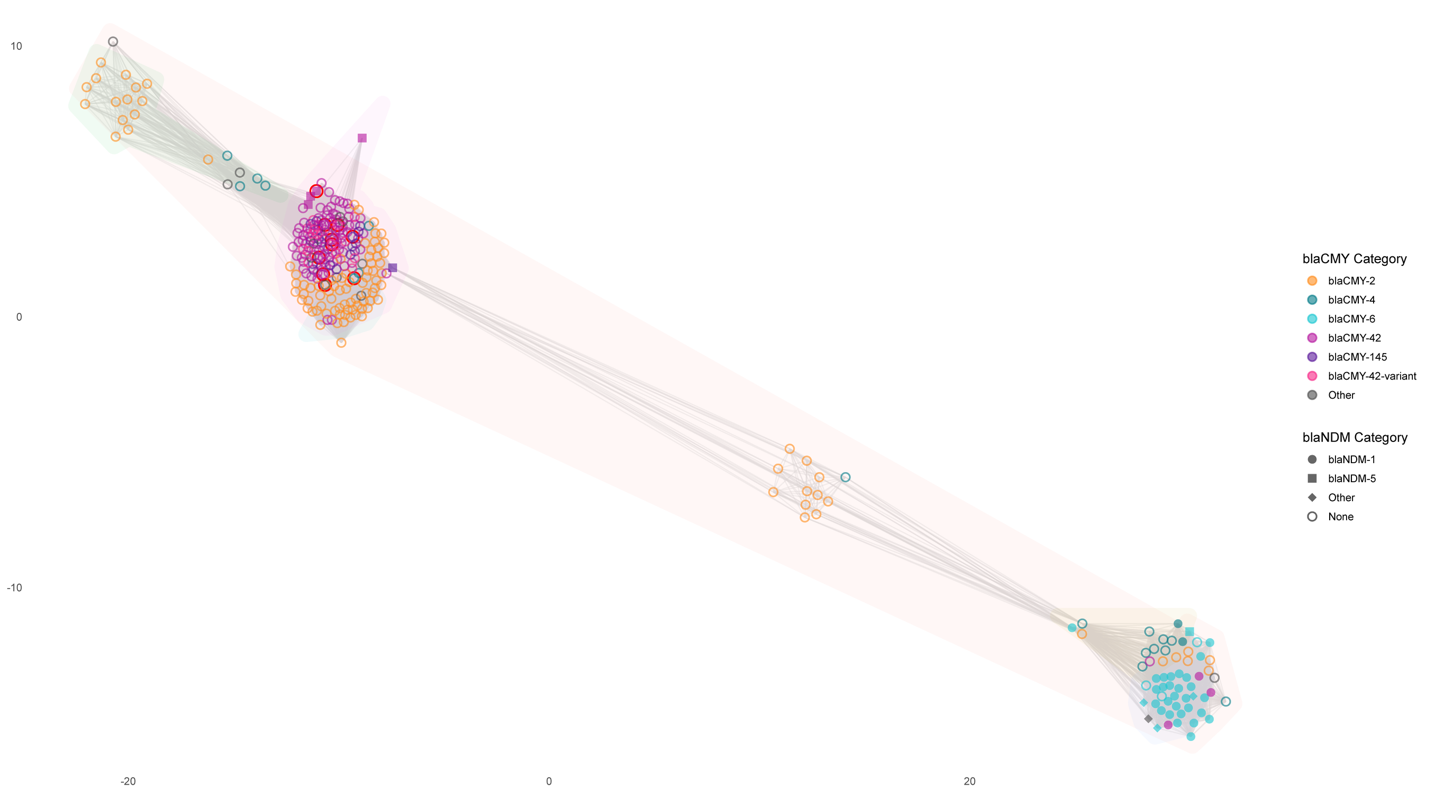

**Fig S3** CMY plasmid network analysis stratified by blaNDM variant carriage and blaCMY variant carriage. Red circles identify this study plasmid cohort.

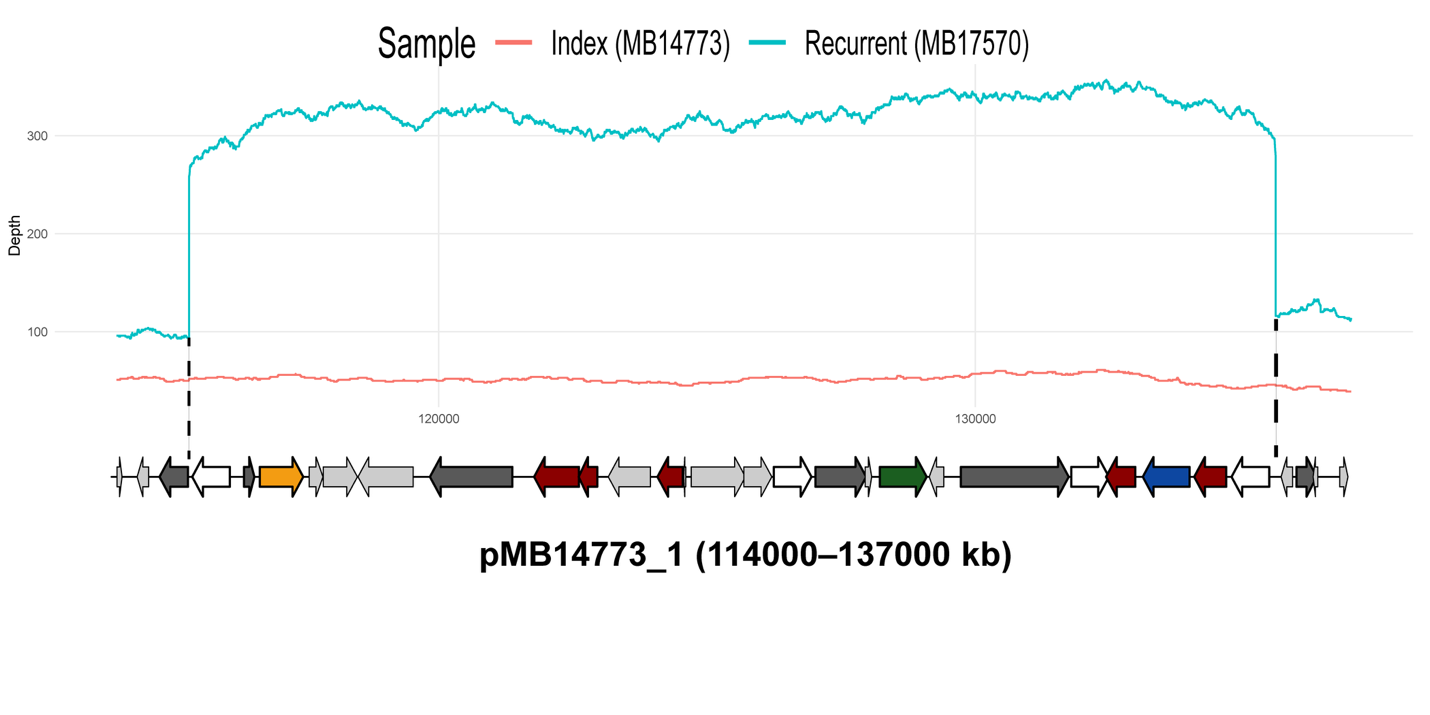

**Fig S4** Read mapping per base coverage depth of pMB14773_1 region (114000 to 137000 kb) for index (red line) vs recurrent (blue line) isolate indicating PCTN-mediated amplification of IS*26* (white arrows) mediated PCTN harboring blaNDM-5 (orange arrows), blaCTX-M-15 (green arrows), blaOXA-1 (blue arrows), and other AMR genes (dark red arrows). Dark gray arrows indicate other transposases present. Dotted vertical black line indicates the boundaries of IS*26* amplification demarcated by the right inverted repeat (IRR) and left inverted repeat (IRL) of cis oriented IS*26* transposase genes reading left to right.
